# Scrutinizing the Spread of Covid-19 in Madagascar

**DOI:** 10.1101/2020.09.27.20202556

**Authors:** S. Narison, S. Maltezos

**Affiliations:** Laboratoire Univers et Particules de Montpellier (LUPM), CNRS-IN2P3, Case 070, Place Eugène Bataillon, 34095 Montpellier, France; Institute of High-Energy Physics of Madagascar (iHEPMAD), University of Ankatso, 101 Antananarivo, Madagascar; National Technical University of Athens, Physics Department, Heroon Polytechniou 9, Zografos GR15780 Athens, Greece

**Keywords:** Covid-19, Epidemic, Pandemic, Infectious disease, Virus spread, Confinement

## Abstract

We scrutinize the evolution of Covid-19 in Madagascar by comparing results from three approaches (cubic polynomial, semi-gaussian and gaussian-like models) which we use to provide an analytical form of the spread of the pandemic. In so doing, we introduce (for the first time) the ratio 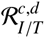 of the cumulative and daily numbers of infected persons over the corresponding one of tests which are expected to be less sensitive to the number of the tests because the credibility of the results based only on the absolute numbers often raises some criticisms. We also give and compare the reproduction number *R*_eff_ from different approaches and with the ones with the ones of some European countries with a small number of population (Greece, Switzerland) and some other African countries. Finally, we show and comment the evolution of the total number of deaths and of the per cent number of cured persons and discuss the performance of the medical care.

## 1. Introduction

In a previous paper [1], we have carefully analyzed step by step the spread of COVID-19 during the two first months of the pandemic until the 48th day (07/05/20), where a sudden jump signaling an eventual second phase of the pandemic has emerged.

In this paper, we pursue this analysis until 12th September which is included in the region after the peak of the second phase.

To avoid some criticisms on the unsufficient number of tests and on the eventual strong dependence of the number of infected persons on the number of tests, we have introduced the cumulative ratio :

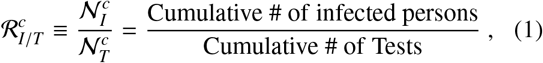

and the corresponding daily ratio :

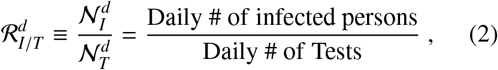

which we shall study, in the following.

We shall attempt to give an analytical form of the different data by comparing the predictions of three models :

– A cubic polynomial which has described successfully the small phase of the pandemic [1] :

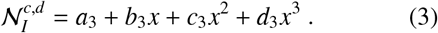
– An hybrid-exponential Model, hereafter named Large Population Semi-Gaussian (LPE-SG) Model [2] (see also [3]):

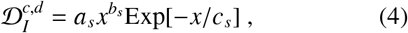

where : *a*_*s*_ ≡ *A* is an arbitrary amplitude, *b*_*s*_ ≡ *n* the degree of the model and *c*_*s*_ ≡ *τ* the mean infection time from which one can deduce the position of the peak *x*_*p*_ = *b*_*s*_ *c*_*s*_.
– A Gaussian-like Model [4]:

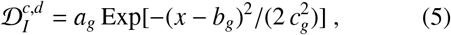

where : *a*_*g*_, *b*_*g*_, *c*_*g*_ are the height, position and width of the Gaussian.

*𝒩*_*I*_ / 𝒟_*I*_ is the cumulative / daily number of infected persons while the coefficients : *a*_*i*_, *b*_*i*_, *c*_*i*_ and *d*_*i*_ will be determined from the fitting procedure.

## 2. Data on the number of infected persons

### • Cumulative numbers

The data from [5–7] are shown in Fig. 1a) for the absolute number of infected persons and in Fig. 1b) for the ratio 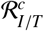 defined in Eq. 1.

**Figure 1:**
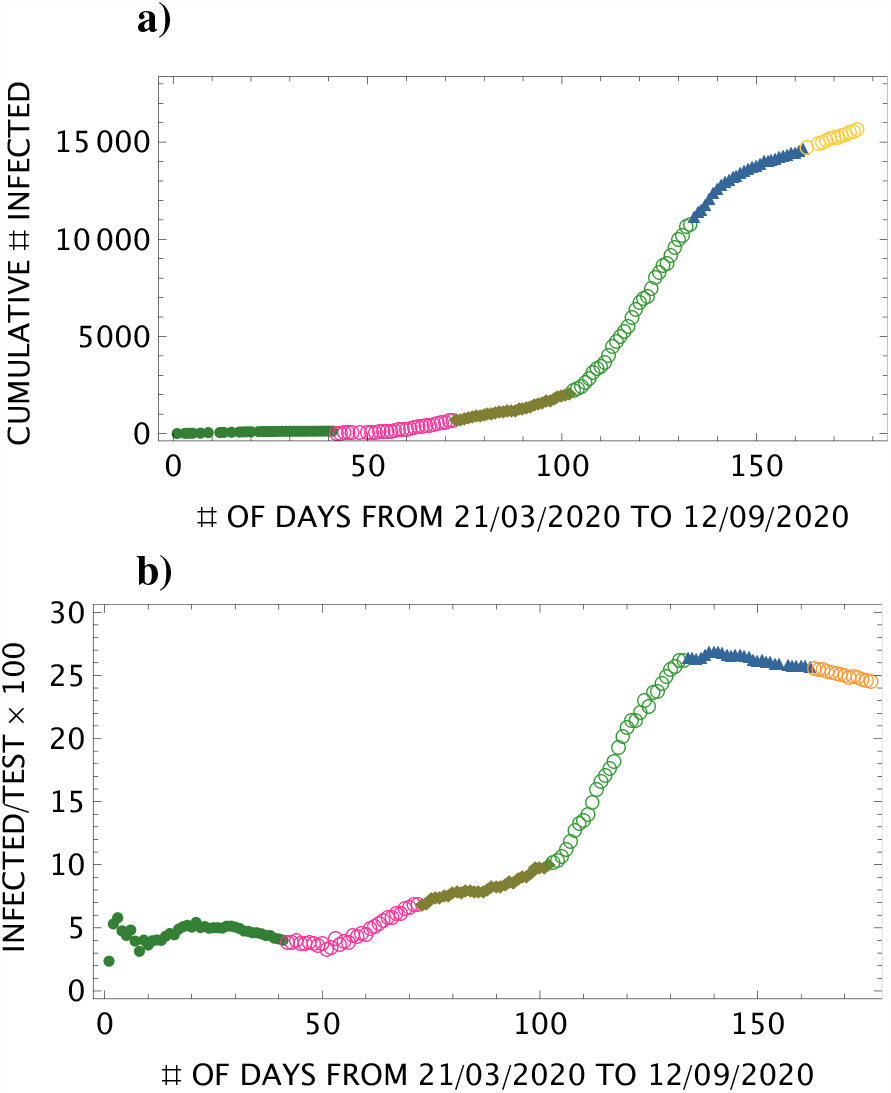
Daily behaviour of the cumulative # of infected persons from 21th March to 12th September : **a)** the 1st month case until the end of april (fully oliva points); May (open pink circle); June (oliva full square); July (open green circle); august (full dark blue triangle); September (open yellow circle); **b)** Same as **a)** but for the cumulative ratio 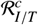 defined in Eq. 1.

One can notice that the shape of the two curves shown in Figs.1 are quite similar expect at the period after the peak of the 2nd phase of the pandemic, where the ratio shows clearly the decline of the pandemic. This (surpising) feature indicates that the number of tests affects only slightly the behaviour of the evolution of the pandemic.

### • Daily number

The daily behaviour of the ratio 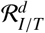 defined in Eq. 2 is shown in Fig. 2 : **a)** The first months behaviour until 7th May; **b)** the second phase behaviour from 7th May until 12th September.

**Figure 2:**
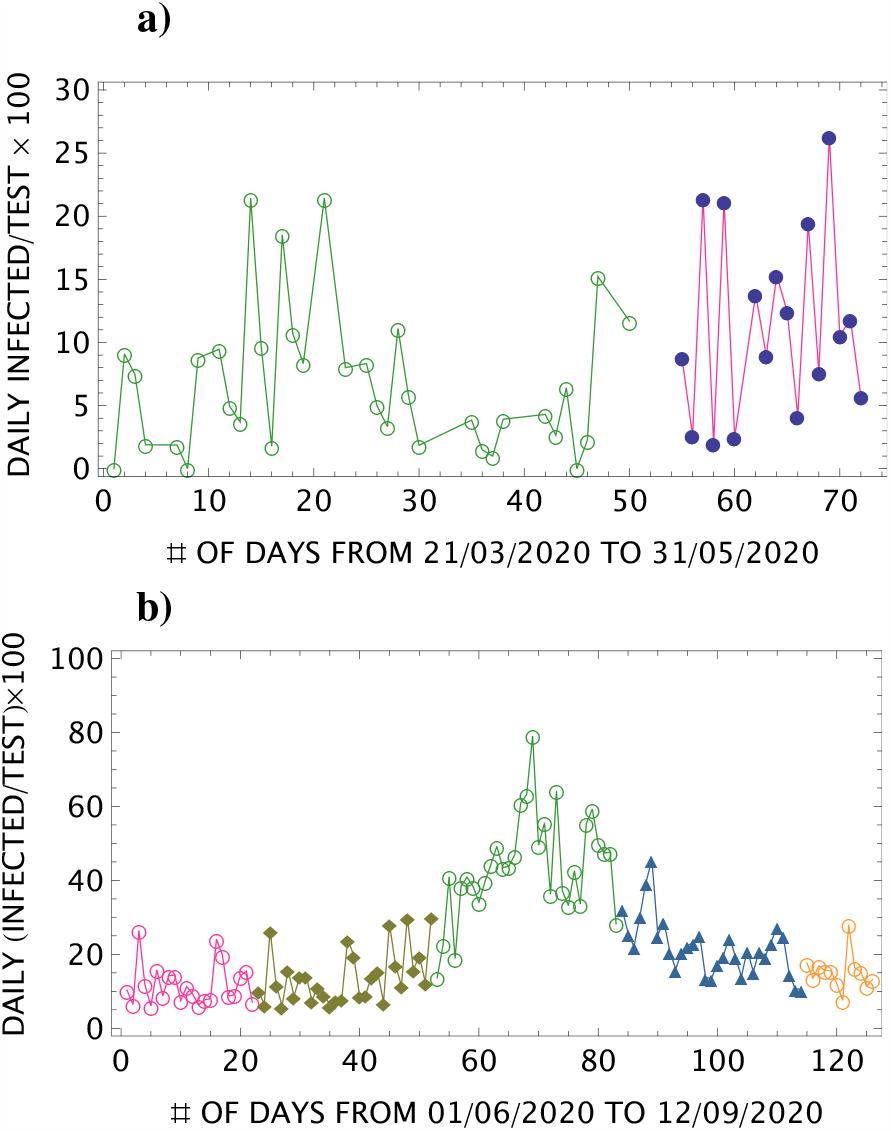
Daily behaviour of the cumulative # of infected persons: **a)** the 1st month case until 7th May (fully oliva points), 7-31th May (open pink circle); **b)** 1-22th June (open pink circle), 23-30th June (oliva full square), July (open green circle), August (full dark blue triangle), 12th September (open yellow circle).

### • Phases of the pandemic

One can notice from the previous figures that there are two phases :

– Small or Epidemic phase until 48 days (7th May).
– Large or Pandemic Phase after 7th May.

We shall see later on that these two phases can be sub-vided into two phases ⊕ a background phase ⊕ from 7th to 31th May.

## 3. The small phase until 07*/*05*/*2020 (48 Days)

### • Absolute cumulative number of infected persons

Detailed analysis and a comparison of different models can be consulted in [1]. One can deduce, from the analysis done there, that the peak (stability of the cumulative number of infected persons) is reached for 25 to 30 days after the beginning of the pandemic (stability). The two models LPE-SG and Gaussian-like models according to the parameters fitted in this paper gives a value around 40 days which is still compatible with the data though in the higher sides.

### • Cumulative ratio 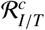

A similar analysis is repeated here but with the ratio of the cumulative number of infected persons over the cumulative number of tests. The results of the fit are given in Fig.3a) where the corresponding best fit parameters are :

**Figure 3:**
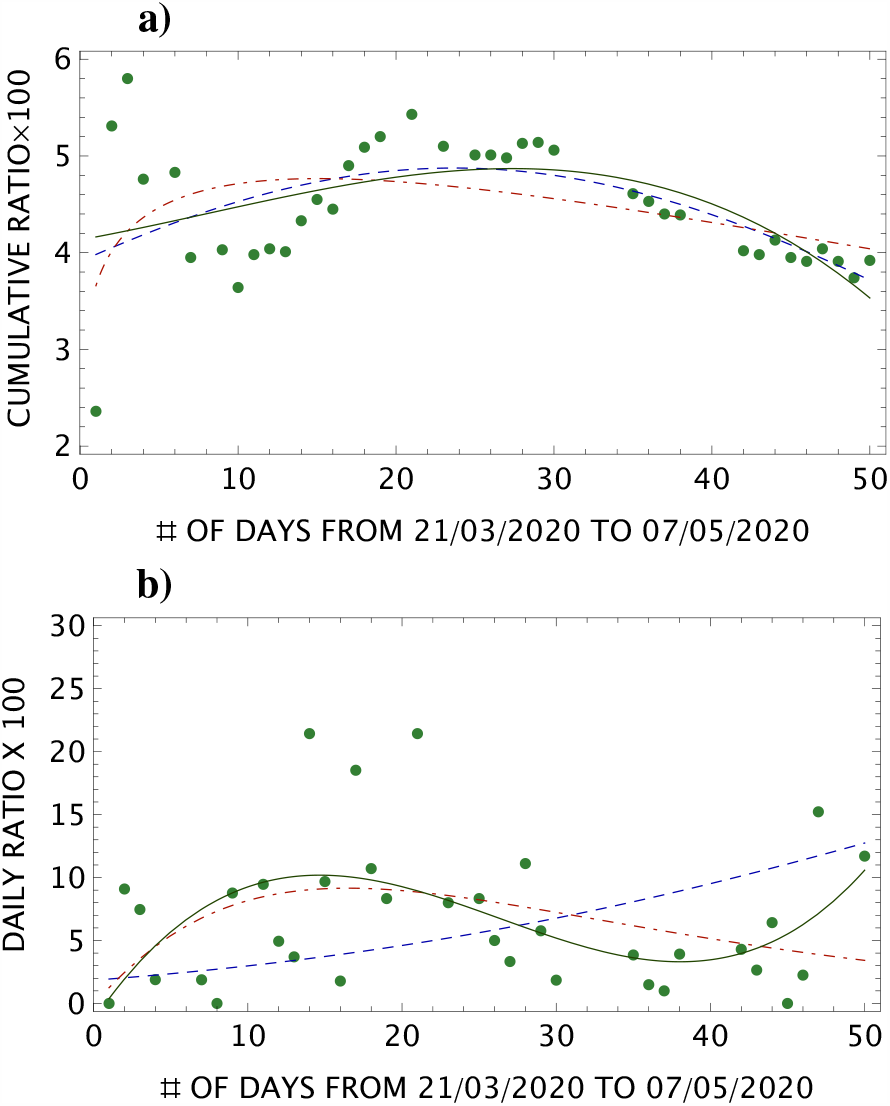
Data and analysis of : **a)** cumulative ratio 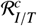 defined in Eq. 1; **b)** daily ratio 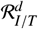 defined in Eq. 2.

– Cubic Polynomial [1]:

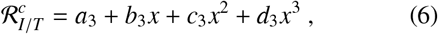

where one finds :

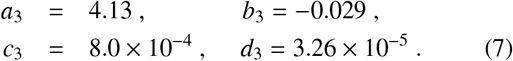
– LPE-SG Model [2]:

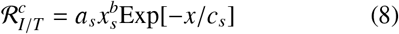

with :

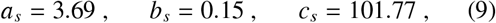
– Gaussian-like model [4]:

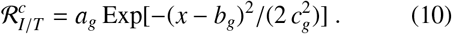

with :

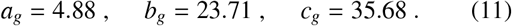

From the previous analysis, one can see that the cubic polynomial finds a maximum peak around the 30th day, the LPE-SG model around 15 days and the Gaussian-like one around 40 days. The results from the cubic polynomial and Gaussian-like model are consistent with the absolute one obtained in [1], while the one from LPE-SG is much lower (perhaps) indicating that the Model is not quite appropriate for analyzing the ratio 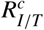.

### • Daily ratio 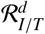

Similar analysis is done for the daily ratio which is shown in Fig. 3. The parameters fit are:

– Cubic Polynomial [1]:

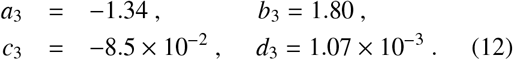
– LPE-SG Model [2]:

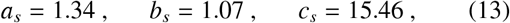
– Gaussian-like model [4]:

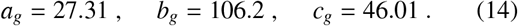

From the previous analysis, one can see that the cubic polynomial and the LPE-SG model find a maximum peak around the (14-17)th day while the Gaussian-like model peak is around 46 days (not shown in the figure).

### • Comments on the results

Comparing the results from the cumulative and daily ratios, one can see that the LPE-SG prediction of the peak position is almost the same (15-17)th day for the cumulative and daily ratios, the cubic polynomial predicts a peak around (14-28)th day and the Gaussin-like around (28-46)th day. One can also notice from Fig. 3b) that the 3 models do not give a good prediction of the intensity of the daily peak unless considering that the three data points shown there around 15-20th days are statistical flucuations. This discrepancy is less pronounced for the cumulative ratio shown in Fig. 3a). Analogous observation on the imprecision of models to predict the intensity of the peak has been discussed in Ref. [8] for the case of the Wuhan pandemic.

## 4. Before the 2nd peak : 07*/*05*/*2020 to 12*/*08*/*2020

### • Absolute cumulative number of infected persons

The result of the analysis is shown in Fig. 4.

**Figure 4:**
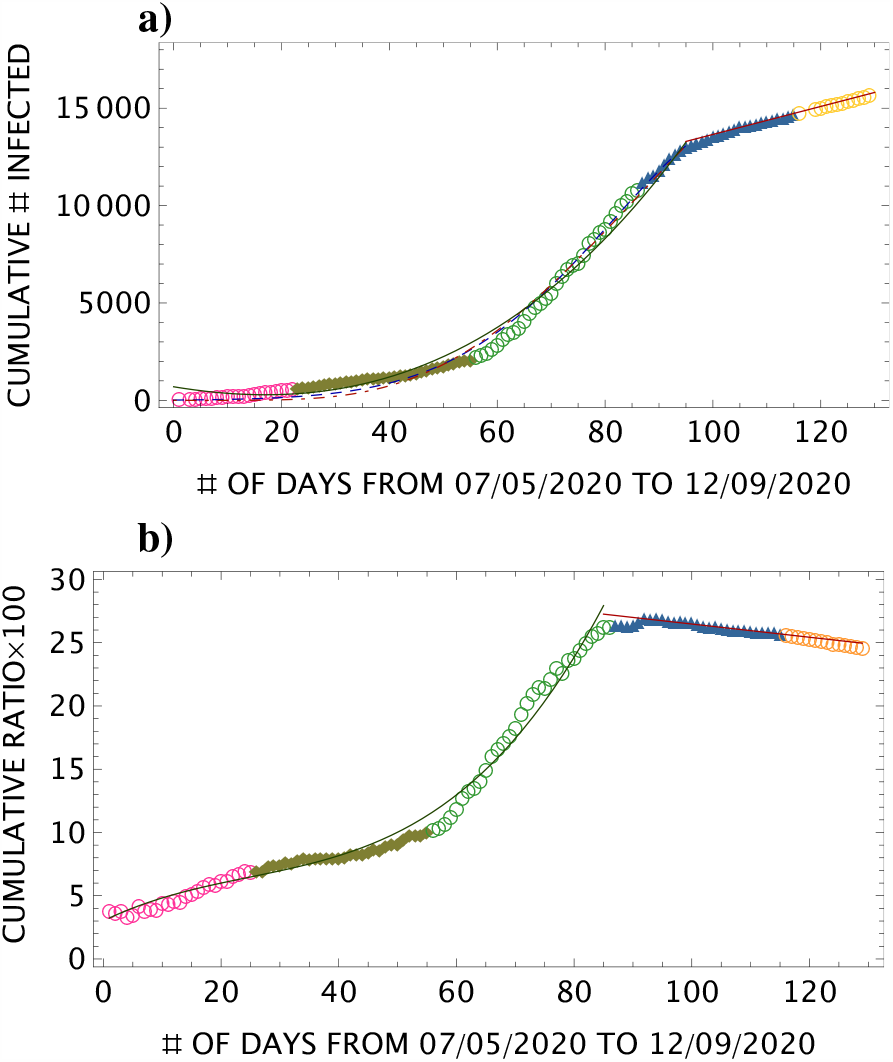
**a)** Fit of the absolute cumulative # of data in Fig.1a) from 50 days to 100 days; **b)** same as in **a)** but for the ratio. The legends are the same as in Fig. 1.

– Cubic Polynomial [1]:

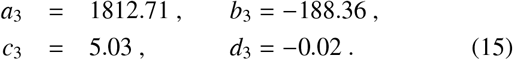
– LPE-SG Model [2]:

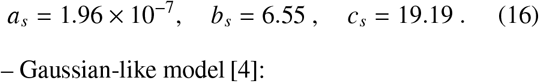
– Gaussian-like model [4]:

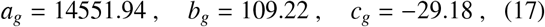

From the previous parameters, one finds that the LPE-SG model finds a peak around *b*_*s*_ × *c*_*s*_ ≈ 126 days, while the Gaussian-like model finds a gaussian centered at 109 days which are compatible. One can notice that the cubic polynomial gives a good fit of the data but cannot predict a peak

### • Ratio ℛ_I/T_ of # Infected persons over the # of Tests

Doing a similar analysis for the ratio ℛ_*I/T*_, we show in Fig. 4 the fit from the cubic polynomial with the parameters:

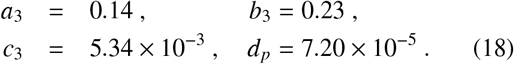

We do not show the fits from the two models LPE-SG and Gaussian-like which are not good and which lead to unrealistic values of the fitted parameters.

One can notice that the fit from the 2 models are less good here than in the absolute case shown in Fig.4.

## 5. After the 2nd Peak of 12*/*08*/*2020

After the peak, one can see that the total number of infected persons increase linearly as :

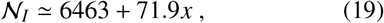

which corresponds to an increase of about 72 infected persons / day, while the ratio ℛ_*I/T*_ decreases slightly as:

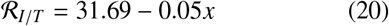

corresponding to - 0.05% infected person per day. It indicates that the cumulative number of infected persons decreases slowly when the cumulative number of tests increases which is a clear signal that the pandemic is decreasing.

## 6. Fit of the 2nd phase from 07*/*05*/*2020 to 28*/*08*/*2020

Here, we redo the analysis but for a global fit of the 2nd phase. The fitted parameters are :

### • Absolute cumulative number of infected persons

The analysis is shown in Fig. 5.

**Figure 5:**
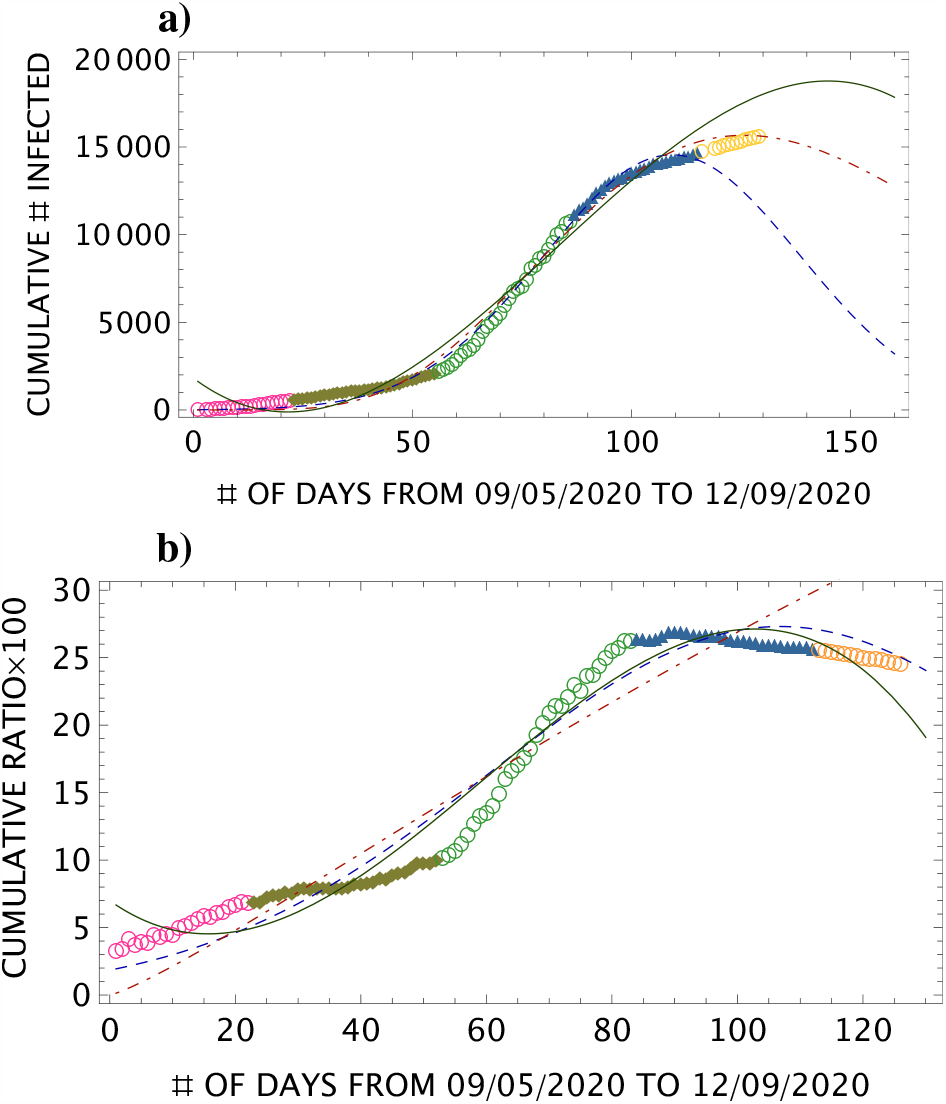
Fit of the data in Fig.1 from 50 days to 130 days: **a)** absolute cumulative # of infected, **b)** cumulative # of infected over cumulative # of tests. Same legends as in Fig. 1. The open yellow circles at the end of the curve are new data not used in the fit. The origin of the axis is taken at the 50th day. The continuous oliva line is the fit from a cubic polynomial. The dashed blue curve is the one from a Gaussian-like model. The dot-dashed red curve is the fir from the LPE-SG model.

– Cubic olynomial:

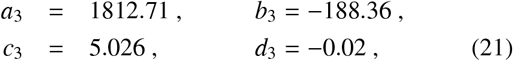
– LPE-SG Model :

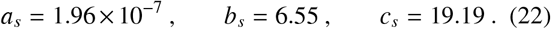
– Gaussian-like Model :

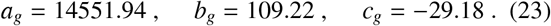

The polynomial fit shows a maximum around 150 days. The LPE-SG Model indicates a peak around : *b*_*s*_ × *c*_*s*_ ≃ 126 days, while the Gaussian is centered at 109 days. These predictions are compatible each others.

### • Cumulative ratio 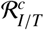

The results from different models are shown in Fig.5 which correspond to the parameters :

– Cubic Polynomial [1]:

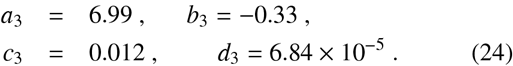
– LPE-SG Model [2]:

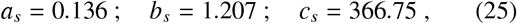
– Gaussian-like model [4]:

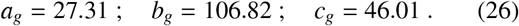

One can deduce from the analysis that the cubic polynomial fit and the Gaussian-like model indicate a peak around (105–107) days i.e on (3–7) th of July, while the values of the LPE-SG fitted parameters are not conclusive when analyzing the ratio 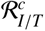.

However, if we start the analysis from 15th June (origin of the x-axis), we obtain a much better fit (see Fig.6) with reasonnable values of the fitted parameters :

**Figure 6:**
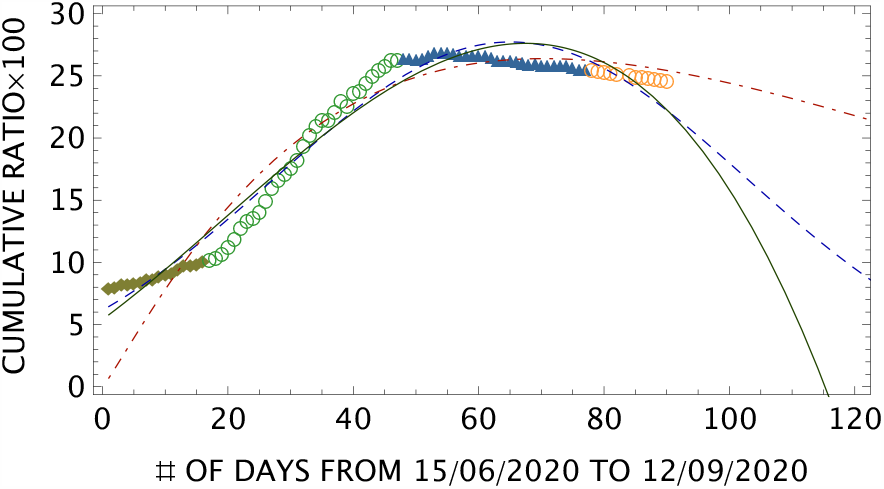
Fit of the data in Fig.1 from 87th day (15th June) : same legends as in Fig. 1. The origin of the axis is taken at the 87th day. The continuous oliva line is the fit from a cubic polynomial. The dashed blue curve is the one from a Gaussian-like model. The dot-dashed red curve is the fit from the LPE-SG model.

– Cubic Polynomial [1]:

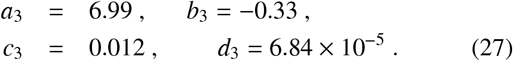
– LPE-SG Model [2]:

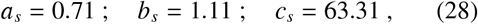
– Gaussian-like model [4]:

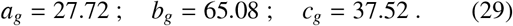

The three appraoches predict a peak around the (65-70)th day i.e around (19-24)th august 2020.

## 7. The reproduction numbers *R*_eff_ of the 1st phase

We analyze these quantities using the definition proposed in [9] based on a SIR model [10], where the discrete instantaneous reproduction number is:

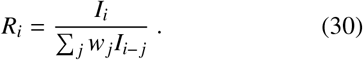

*I*_*i*_ is the number of infected persons at a time *t*_*i*_ which follows a Poisson distribution; *w*_*i*_ is the measures of infectiousness with the properties : 0 ≤ *w*_*i*_ ≤ 1 and ∑ *w*_*i*_ = 1.

### • Lower bound on the initial reproduction number R_0_

We attempt to provide a lower bound on *R*_0_ by using the data reported on the first four days from 19/03/20 to 22/03/2020 where the daily infections are respectively 3,0,0 and 9. Using these data and the properties of *w*_*i*_, one can deduce :

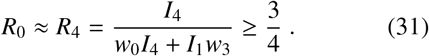

The bound is rather weak but may reflect the epidemic (not yet a pandemic) nature of the disease. It may be also useful to improve the accuracy of predictions of *R*_0_ from some other models.

### • Value of R_0_

Considering that the result from the 1st week is statistically poor, one can deduce from Fig. 7 the rough estimate from the beginning of the 2nd week :

**Figure 7:**
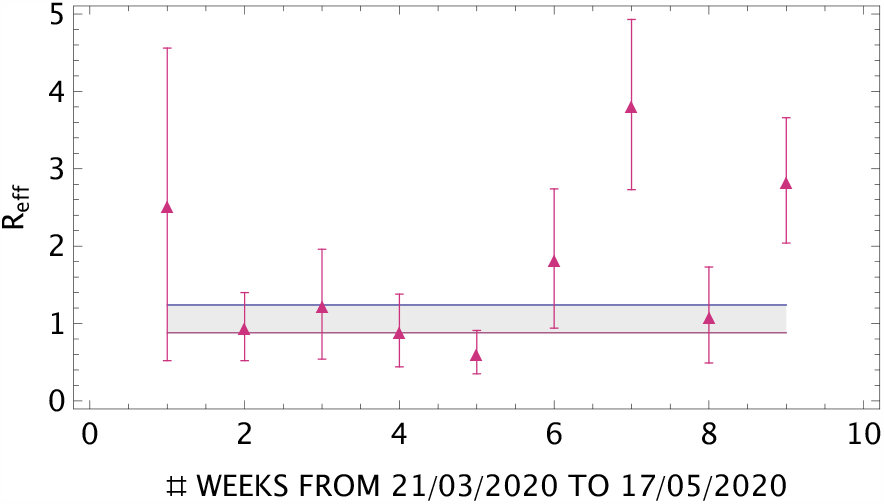
Weekly behaviour of the reproduction number *R*_eff_ of infected persons until 17th May. The dashed region is a tentative weighted average.

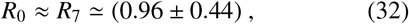

### • The effective reproduction number R_eff_

Using the previous SIR-like model, we attempt to extract the instantaneous / effective reproduction number *R*_eff_ for an interval period of about one week. We show the results in Fig.7. The large error bars take into account the low and high values of the predictions obtained in [7]. In the dashed region, we show the tentative weighted average :

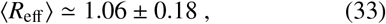

where its relatively low value may indicate that this first phase is still an epidemic but not yet a pandemic period.

## 8. *R*_eff_ from the LPE-SG model

### • The three phases

We have noticed in the previous analysis that the LPE-SG model fitted parameters are very sensitive to the starting / ending points of the data. The reason is that the model is asymmetric contrary to the gaussian-like model and the cubic polynomial. Therefore, for better exploiting this model, it is more convenient to divide the total period of the pandemic into 3 phases (see Fig. 8) :

**Figure 8:**
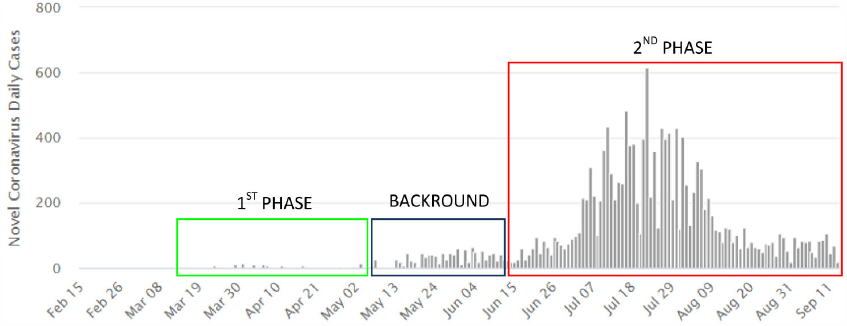
Different phases of Covid-19 in Madagascar for the daily number of infected persons.

– The 1st phase for the first 2 months until 2nd May (epidemic).
– The phase between 2nd May to 15th June (background), which is below the pandemic threshold where the data fluctuate eventually showing the beginning of the pandemic. It can be considered as a continuation of the 1st phase.
– The 2nd phase beyond 15th June (real pandemic).

We show in Fig.9 the predictions of the LPE-SG model corresponding to the parameters :

**Figure 9:**
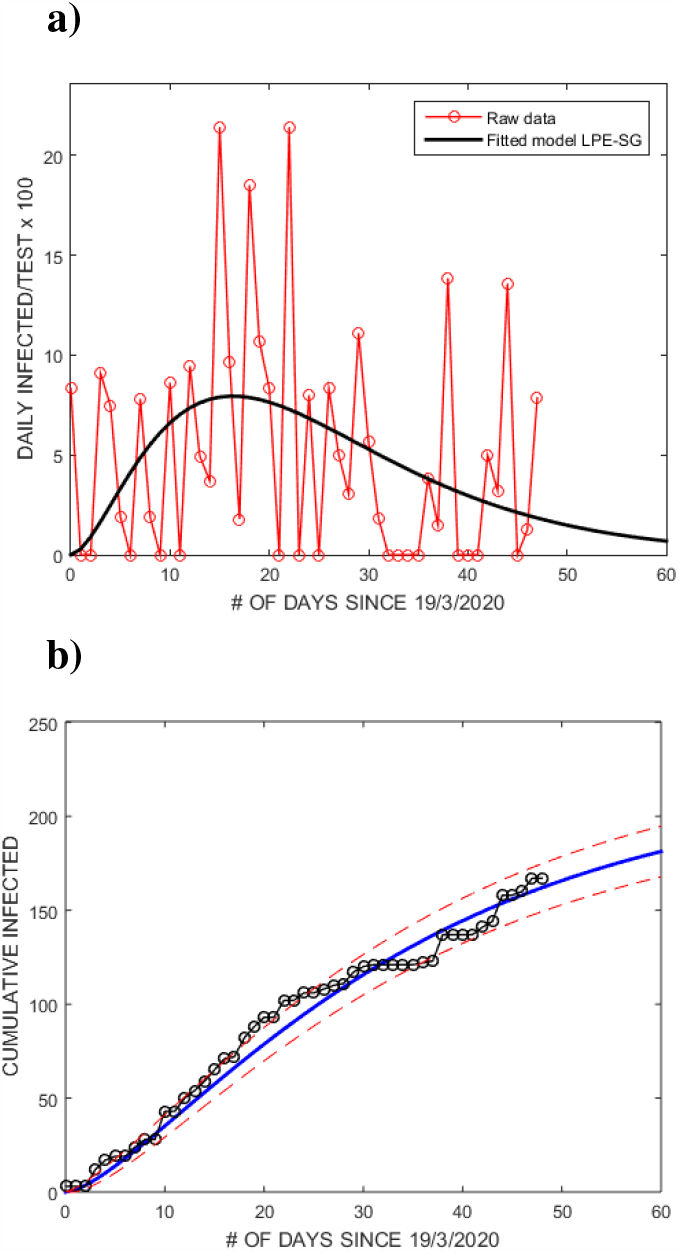
Fit of the daily **a)** and cumulated **b)** reported data for infected persons using LPE-SG model for the 1st phase from 19th March until 2nd May.

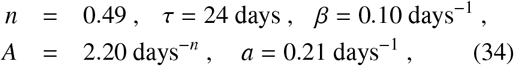

which give :

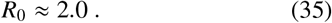

### • R_0_ of the 1st phase

The position of the peak around 12 days is consistent with the one obtained in Fig. 3 by a direct fitting of the data from which one can deduce a more precise value of *R*_0_ than in the previous Eq. 35 :

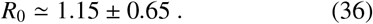

This value is comparable with the one from SIR-like model in Eq. 32.

### • R_eff_ of the 1st phase

We study in Fig. 10 the behaviour of *R*_eff_ using the daily ratio of corrected data.

**Figure 10:**
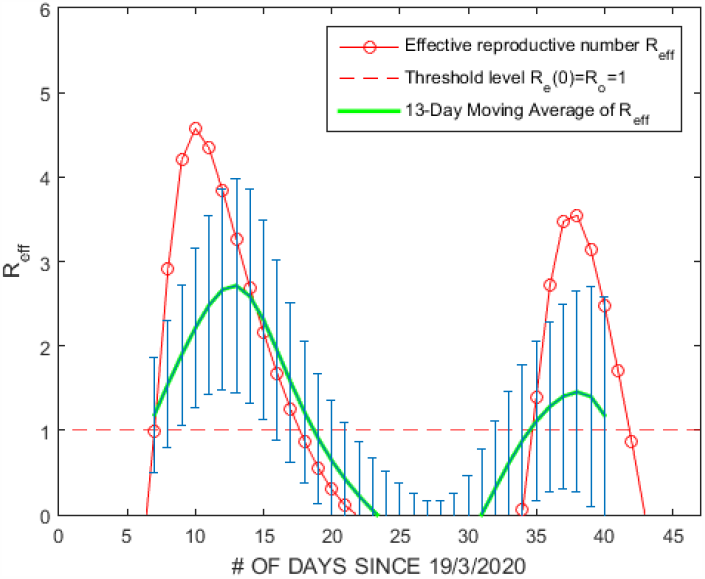
*R*_eff_ of the first phase using LPE-SG model from 29th March to 7th May. The green curve is the moving average of *R*_eff_ every 13 days.

### • R_0_ of the 2nd phase

We redo the analysis but for the 2nd phase from 15th June. Using the corrected daily tests, we show in Fig.11 the predictions of the LPE-SG model corresponding to the parameters :

**Figure 11:**
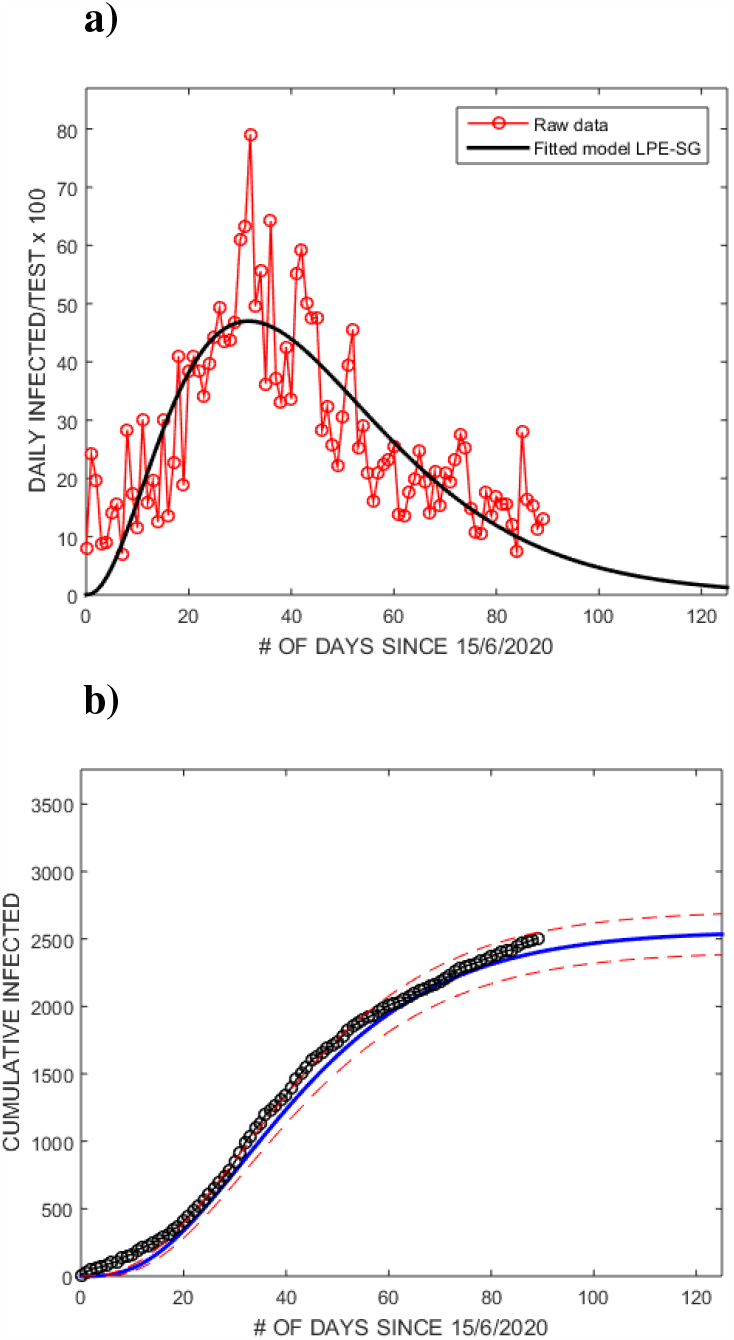
Fit of the daily **a)** and cumulated **b)** reported data for infected persons using LPE-SG model from 15th June to 12th September.

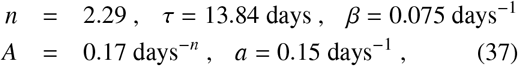

which give :

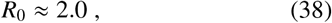

and a peak around 34 days from 15th June i.e 121 days in total (19th July). However, the peak is obtained earlier than in Fig. 6 from a direct fit of the data and using different models.

### • R_eff_ of the 2nd phase

– To estimate *R*_eff_, we use its definition according to the SIR model and obtain the following useful expression:

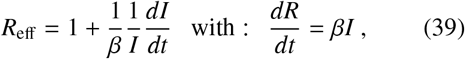

where *β* is the recovering rate which is the inverse of the recovering time symbolized with *τ*_*R*_. This *β*-parameter has to be determined, either from published results or by fit using the LPE-SG model via the *τ*_*R*_ parameter. However, apart of its stochastic behavior, *β* is likely a function of time for a particular country.
– Starting with a well adjusted value *β* = 0.075 days^-1^, we show in Fig. 12a) the raw data of daily infected *I*(*t*) persons (red open connected circles) and the one of cured / recoverded (green open connected circles) as function of time. The same data are shown in Fig. 12b) but after a low pass filter obtained using Matlab alogrithm which is expected to reduce the statistical (stochastic) variations of the data. The full blue connected circles show the prediction of *R*_eff_ using Eq. 39. It is important to observe that the data of *R*(*t*) the green one after the 40th day diverges from the expected one based on the theory of SIR. This underestimated values can be explained either due to the way of counting the recovered (cured) people, or the rate *β* is a function of time and has been changed (decreased) suddenly for unknown reasons, leading to a serious discrepancy from the basic SIR model.

**Figure 12:**
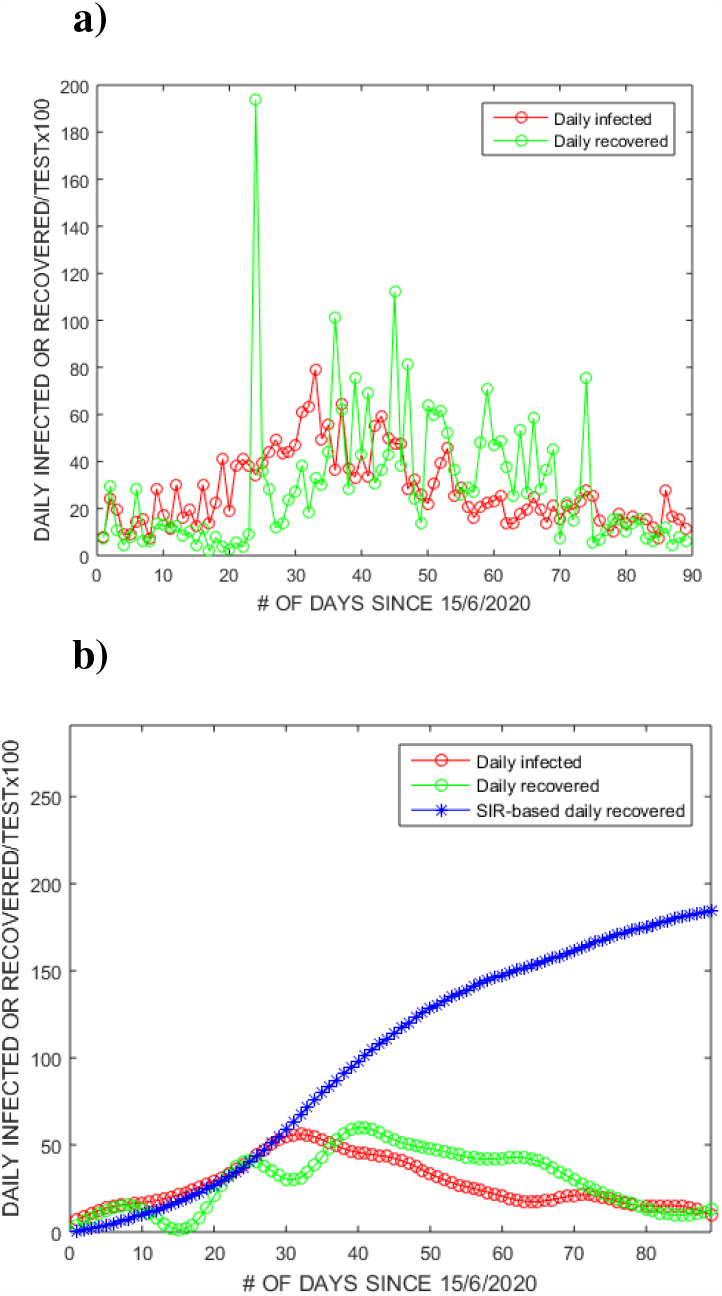
The daily reported raw data **a)** and the smoothed ones by using digital filter **b)**, included the SIR-based daily recovered, from 15th June to 12th September.
– We show in Fig. 13 the predicted values of *R*_eff_ as function of the number of days using the daily infected data (open red circles) given in Fig. 12. One can notice like in the previous sections that the analysis from the LPE-SG models are quite sensitive to the starting and ending points where the results are very inaccurate. Around (35-45) days, the values of *R*_eff_ pass below one and reach a minimum around the 60th day. However, the increases above the 60th day which can be due to the non-monotonic decrease of the data beyond this date (Fig. 12) deserves more attention.

**Figure 13:**
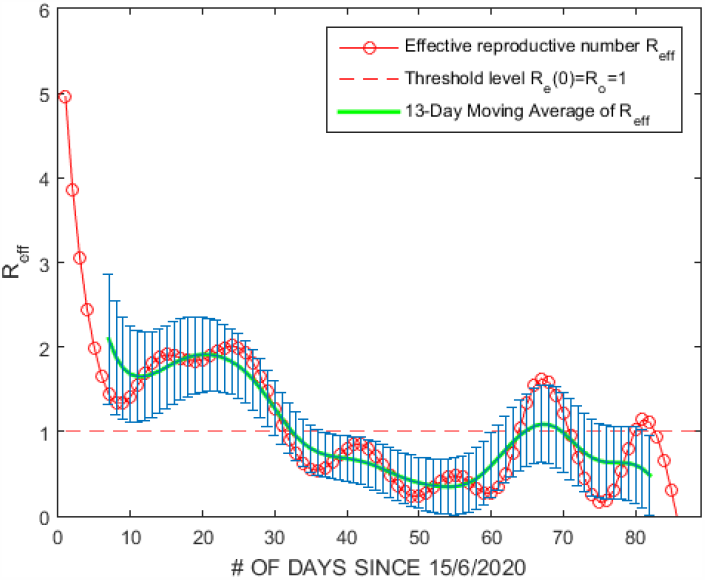
*R*_eff_ of the second phase using LPE-SG model from 15th June to 12th September. The green curve is the moving average of *R*_eff_ every 13 days.

## 9. Comparison of *R*_eff_ from different models

### • The first phase

– We compare the results of *R*_eff_ with the ones from Imperial College [11] shown in Fig. 14. One can notice a good agreement of the SIR-like model predictions in Fig. 7 (2nd column) within the errors with the ones from [11] (3rd column):

**Figure 14:**
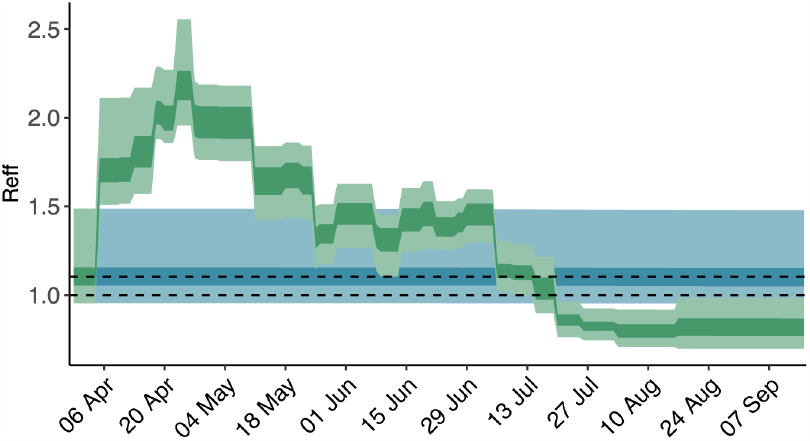
*R*_eff_ of the second phase from 15th June to 12th September.

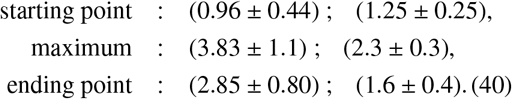
– One can also see a good agreement of the *R*_0_ value from the SIR-like model in Eq. 32 with the one from the the LPE-SG model in Eqs. 35 and 36.

### • The second phase

– For the 2nd phase shown in Fig. 13 from the LPE-SG model, one can notice a good agreement with the ones from Imperial College in Fig. 14 [11]. We deduce from Fig. 13:

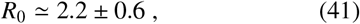

for the 2nd phase.
– We consider the case of Greece and Switzerland as these countries has relatively few numbers of population (≈ 10.42 billion for Greece and 8.57 billion for Swizerland) which may be comparable with the one of Madagascar (≈ 27.69 billion). We show the value of *R*_eff_ obtained using the same LPE-SG model in Fig. 15 where one can notice that the value of *R*_eff_ as a function of time are lower in Madagascar than that in Greece (GR) at the first outbreak of Covid-19 :

**Figure 15:**
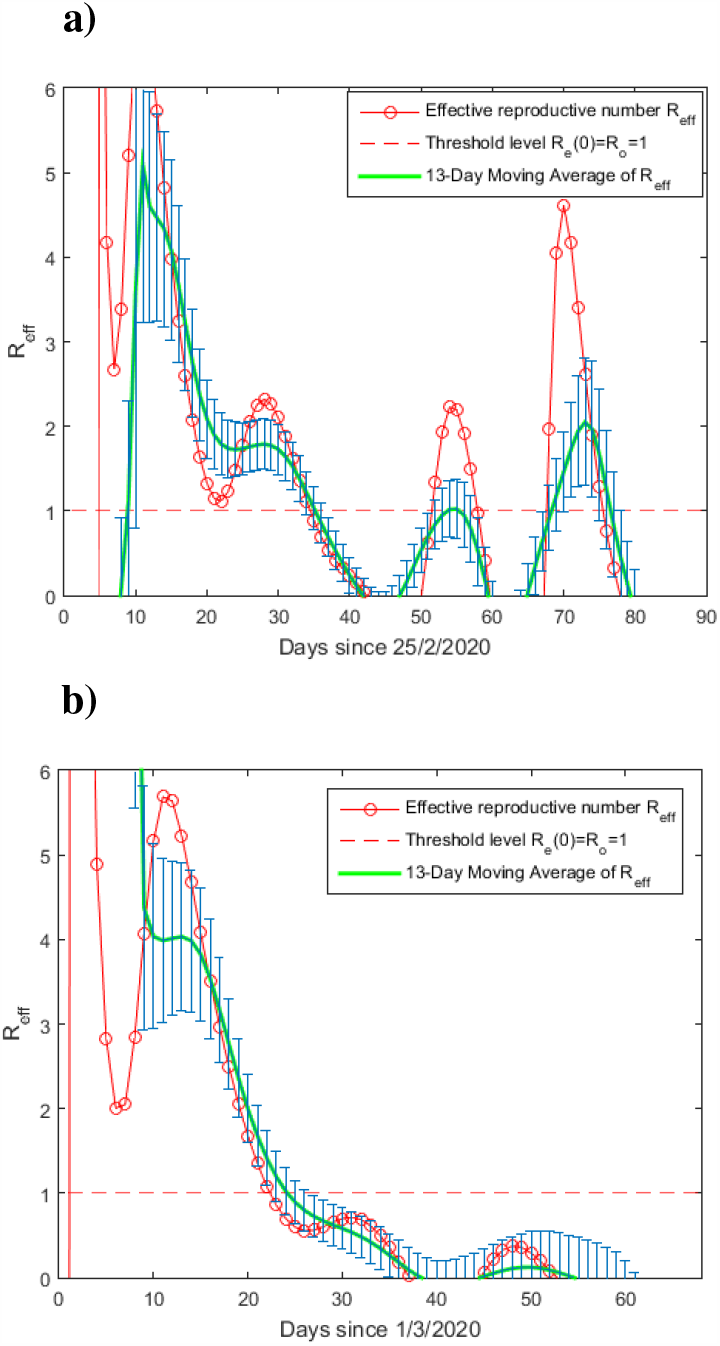
*R*_eff_ of the first outbreak using the LPE-SG model from 25th February for : **a)** Greece ; **b)** Switzerland. The green curve is the moving average of *R*_eff_ every 13 days.

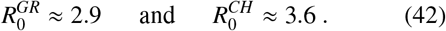
– We show in Fig. 16 the results for *R*_eff_ for different African countries including Madagascar from Imperial College studies where one can notice that the prediction :

**Figure 16:**
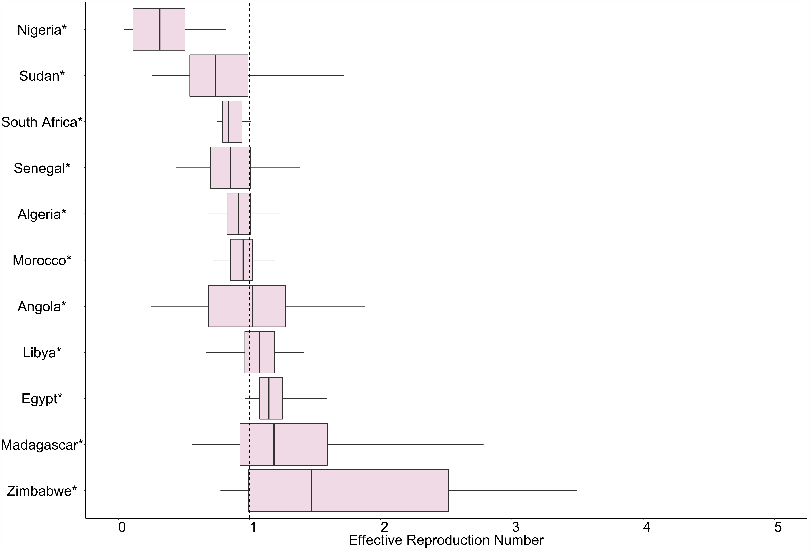
*R*_eff_ from recent analysis of different African countries from Imperial College studies.

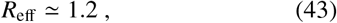

agrees with our tentative average ⟨*R*_eff_⟩ ≃ (1.06 ± 0.18) of the first phase in Eq. 33 from a SIR-like model but smaller than the one from the LPE-SG model of about 2. This difference indicates the difficulty of each model to provide an accurate prediction, a point discussed in [8].

## 10. Absolute Number of Deaths until 12*/*09*/*20

The data on the absolute # of deaths are shown in Fig. 17. A cubic polynomial fit which smears the data is also shown. The parameters fit are :

**Figure 17:**
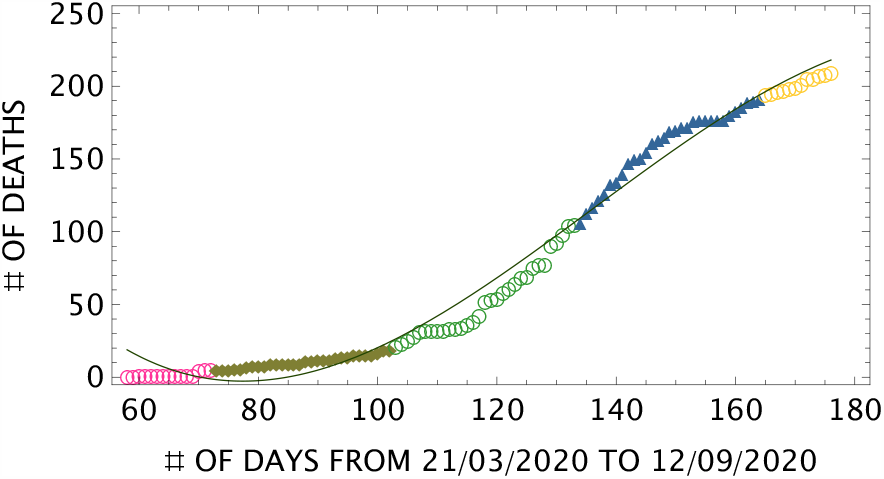
Daily behaviour of the cumulative number of deaths until 12th September. The oliva curve is a smearing of the data with a cubic polynomial.

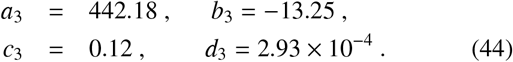

One can notice that the (officially declared) absolute cumulative number of deaths are hopefully small. However, the unofficial observation of the population does not support this small number and needs more clarifications.

## 11. Per cent Number of Cured Persons until 12*/*09*/*20

We show in Fig. 18 the per cent cumulative number of cured persons relative to the one of the infected persons. Looking at the behaviour of the number of cured persons which reflects the performance of the medical cares, one can notice that :

**Figure 18:**
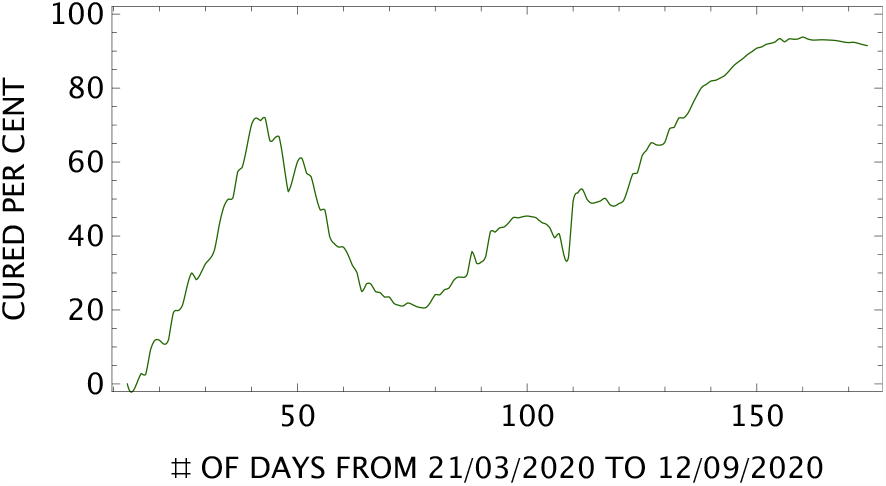
Daily behaviour of the per cent cumulative number of cured persons until 12th September.

- On the first 48th day, the medical care performance was quite good due mainly to the few numbers of infected persons.
- On the 48th day when the new pandemic phase has started and where the number of infected persons has jumped, the medical care performance has decreased until the 80th day. It has indicated the limitation of the hospital equipment and the way to face the pandemics. Notice that the miraculous Covid-organics (CVO) treatment using artemisia leaves mixed with some other traditional plants (white eucalyptus and ravintsara leaves) to cure the pandemic has been officially announced with great fanfare by the Republic President on 29th april was not helpful and inefficient.
- From the 80th day (8th June), the care performance started to increase and becomes excellent after the 150th day just beyond the peak for infected persons around the 110 days (8th July). Many possible effects may have contributed to this success such as the:
  – Uses of the Prof. Didier Raoult protocol based on hydroxychloroquine and azythromycine.
  – More strict sanitary measures (respect of social distancing, masks,…) including confinement.
  – Obtention of more medical equipment thanks to the call of the former health minister,…
  – Relative decrease of the number of infected persons when approaching the peak.

## 12. Summary and conclusions

We have scrutinized the spread of COVID-19 in Madagascar from its beginning to the present day (12th September 2020) where the work has been finalized. We summarize our analysis below :

- We have introduced the ratio 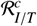 which is the ratio of cumulative number of infected persons over the total number of tests and the analogue daily ratio 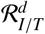 in order to avoid the dependence of the results on the number of tests, as the existing analysis based on the absolute numbers of the infected persons is often criticized on its dependence on the number of tests. However, a comparison of the results from the absolute numbers with the one from the ratio (hopefully and surprisingly) shows that the results are (almost) independent on the number of tests.
- We have also shown that the COVID-19 in Madagascar can be split into three phases : the small phase until 43th days called epidemic where the peak is reached around the (15-28)th day, the transition background phase from 7th May to 15th June and the large phase (real pandemic) beyond 15th June where the peak is reached around the end of august 2020. We have noticed that the gaussian-like and the polynomial fit is not sensitive to the background effect while the LPE-SG does at the starting and ending points of the analysis.
- We have performed our analysis using three independent approaches : a simple cubic polynomial fit, a semi-Gaussian and Gaussian-like models. All aproaches give (almost) consistent predictions of the peak for the absolute cumulative number of infected persons, while for the LPE-SG model to give realistic predictions for the ratio ℛ_*I/T*_, the analysis should be done in the large second phase (3rd period).
- We have used a SIR-like and the LPE-SG models to predict the reproduction numbers *R*_*e f f*_ where we found that the results present large uncertainties both for the small first and large second phases. The difficulty to reach accurate predictions for different models have been discussed in [8]. However, we found that the value of *R*_0_ is relatively low explaining the small number of the infected persons.
- As the pandemic is approaching the ending phase, we do not expect that the results obtained in this paper until 12th september will be affected by the new results not included in the analysis. We expect that the evolution of the cumulative number of infected persons will follow the straightline predicted in Fig. 4.
- The official low number of declared deaths is questionable because it looks to be unrealistic compared to the relative anomalous increase of the observed death number during this pandemic period.
- Our analysis on the evolution of the number of per cent of cured persons shows that the medical care is relatively robust despite the few equipments at disposal. Some factors as the eventual origin of this success have been enumerated previously. However, despite this success, the social and economical conditions of the population induced by ths pandemic are enormous. The often raised “big question” without any clear and convincing answer from the government is the opaque management of the international donated funds dedicated to save the present social and economical situations caused by this pandemic.
- One can finally noticed that during the completion of this work, a new cluster of the pandemic has been detected in the north of Madagascar especially in the small tourstic island of Nosy-Be which may have been induced after the international opening of the airport for tourists.

## Data Availability

The data is available from WHO's site

## Notes

### Competing Interest Statement

The authors have declared no competing interest.

### Clinical Trial

This work reports on data analysis of the spread of Covid-19 and does not need a trial ID

### Funding Statement

This work has not received any fundings

### Author Declarations

This work is a theoretical analysis of the data of the spread of Covid-19 and does not need an IRB number

